# Development of an experimental human blood-stage model for studying *Plasmodium knowlesi*

**DOI:** 10.1101/2025.09.07.25334506

**Authors:** Jeremy SE Gower, Rebecca Webster, Jacob AF Westaway, Jacob A Tickner, Claire YT Wang, Jenny M Peters, Dean Andrew, Angelica F Tan, Lachlan Webb, Adam J Potter, Ria Woo, Susan Mathison, Nischal Sahai, Nanhua Chen, Qin Cheng, Michelle J Boyle, Nicholas M Anstey, Matthew J Grigg, John Woodford, Helen E Jennings, Fiona H Amante, Bridget E Barber

## Abstract

**Background:** The zoonotic parasite *Plasmodium knowlesi* is an increasing cause of human malaria in Southeast Asia. We aimed to develop a *P. knowlesi* induced blood-stage malaria (IBSM) model to enable further study of parasite biology, host responses to infection, and activity of antimalarial treatments.

**Methods:** We manufactured and characterised a *P. knowlesi* parasite bank using the *P. knowlesi* YH1 strain adapted to grow in human serum. This *P. knowlesi* bank was evaluated in an IBSM study involving four healthy adults intravenously inoculated with *P. knowlesi*-infected erythrocytes using a dose escalation strategy (10-fold increases from ~2,800 viable parasites). Parasitemia was monitored by qPCR. All participants received curative treatment with artemether-lumefantrine. Endpoints included safety and infectivity of the *P. knowlesi* parasite bank.

**Findings:** Four participants were inoculated. Two participants became infected. Participant two (administered ~28,000 viable parasites) had detectable parasites (~6 parasites/mL) immediately prior to protocol-defined treatment on day 21. Participant three (administered ~280,000 viable parasites) developed detectable parasites on day 17, peaking at 202 parasites/mL just after protocol-defined treatment on day 24. The parasite multiplication rate over 32 hours (PMR_32_) was 2·33 (95% CI 1·66 – 3·25). Adverse events were mostly mild and unrelated to the challenge agent.

**Interpretation:** A *P. knowlesi* YH1-HS parasite bank suitable for use in human volunteers was successfully manufactured; however, infectivity was suboptimal. Further optimisation of the *P. knowlesi* parasite bank and/or study design will be required to improve infectivity.

**Funding:** National Health and Medical Research Council of Australia and the Royal Brisbane and Women’s Hospital Foundation.

**Research in context:** *Evidence before this study:* Several studies conducted during the 1930s through to the 1960s report the results of *P. knowlesi* human infection studies. This includes the first experimental infection of three individuals in 1931, followed by a number of studies reporting the use of *P. knowlesi* as malariotherapy for neurosyphilis. Several additional studies involving the experimental infection of humans with *P. knowlesi* were conducted in the 1960s, following the first reported case of naturally acquired *P. knowlesi* malaria. No *P. knowlesi* human infection studies have been conducted since the 1960s.

*Added value of this study:* In this study we developed a Good Manufacturing Practice (GMP)-compliant *P. knowlesi* parasite bank suitable for use in human volunteers. This involved adapting the *P. knowlesi* YH1 strain to growth in human serum, resulting in the development of a *P. knowlesi* line (YH1-HS) which can be readily cultured under the same conditions routinely used for *P. falciparum*. We next tested the *P. knowlesi* YH1-HS parasite bank in four healthy volunteers using a dose escalation study. Two participants became infected.

*Implications of all the available evidence:* The development of a reliable and reproducible *P. knowlesi* volunteer infection study would be an important advance for *P. knowlesi* research. While this study resulted in the development of the first GMP-compliant *P. knowlesi* parasite bank, infectivity was suboptimal, suggesting that further optimisation of the *P. knowlesi* parasite bank and/or study design will be required, as well as further investigation into host and/or parasite factors that may impact infectivity.

## Introduction

The monkey malaria parasite *Plasmodium knowlesi* has emerged as a major public health problem in Southeast Asia, with 3290 cases and 14 deaths in 2023.^1^ The majority of *P. knowlesi* cases occur in Malaysia, where *P. falciparum* and *P. vivax* have been eliminated; however, substantial increases have also been reported in neighbouring Thailand and Indonesia.^1^ *P. knowlesi* cannot be reliably distinguished from other *Plasmodium* species by microscopy,^2^ and hence incidence across Southeast Asia is likely underestimated. With its zoonotic reservoir sustaining transmission, *P. knowlesi* represents a major challenge for malaria elimination in the region. Improved understanding of *P. knowlesi*, together with capacity to evaluate antimalarial therapeutics, is critical for ensuring preparedness for an ongoing increase in this zoonotic disease.

Malaria volunteer infection studies (VIS) have proven to be an important tool for studying *Plasmodium* parasites.^3,4^ In addition to allowing efficient evaluation of antimalarial drugs and vaccines, these studies also provide a unique opportunity to evaluate parasite biology *in vivo*, as well as enabling detailed studies of host responses to infection. For *P. falciparum* in particular, induced blood stage malaria (IBSM)-VIS have played a major role in expediting development of antimalarial drugs,^5–8^ and have provided insights into pathophysiological and immune mechanisms in early disease.^9–11^ More recently, IBSM models have also been developed for *P. vivax*^12^ and *P. malariae.*^13^

Substantial experience with *P. knowlesi* human infection studies was obtained prior to the 1960s, particularly through the use of *P. knowlesi* as malariotherapy for neurosyphilis during the 1930s to 1950s.^14–18^ A small number of *P. knowlesi* human infection studies were also conducted following the first reported naturally-acquired human case of *P. knowlesi* in 1965; however, no further *P. knowlesi* VIS have been conducted since this time.^19^

The conduct of IBSM-VIS require access to a suitable malaria challenge agent.^20,21^ Previously, blood-stage parasite banks have been generated by collecting blood from donors deliberately infected with *Plasmodium* strains,^6^ travellers who have acquired malaria naturally while visiting malaria endemic regions^12,13^, or, more recently, through the *in vitro* expansion of laboratory-adapted strains under Good Manufacturing Practice (GMP) conditions.^20–22^ Here we report the manufacture of a GMP-compliant *P. knowlesi* parasite bank utilising the laboratory-adapted *P. knowlesi* YH1 strain, which had previously been adapted for long-term *in vitro* culture in human red blood cells (RBCs) and bovine serum albumin,^23^ and which we further adapted to grow in human serum. We also report the testing of this *P. knowlesi* parasite bank in a pilot IBSM study to evaluate safety and infectivity.

## Methods

### Ethics statement

The study was conducted in accordance with the Declaration of Helsinki and the International Committee of Harmonisation of Good Clinical Practice guidelines. The study was approved by the Human Research Ethics Committees of QIMR Berghofer (P3928) and Lifeblood (2023#42), and registered with the Australian New Zealand Clinical Trials registry (ACTRN12623001326684p). All participants gave written informed consent before enrolment.

### Production of the *P. knowlesi* YH1 pre-seed and seed banks

The *P. knowlesi* YH1 strain, originally generated by Lim et al,^23^ was adapted to grow in media supplemented with only heat-inactivated (HI) human serum to meet GMP requirements^24^ (**Figure 1**), generating the “YH1-HS” parasite line. First, one vial of YH1 was thawed according to methods previously described^25^ and cultured in RPMI 1640 supplemented with 0·5% albumin and hypoxanthine thymidine (HT; 0·1 mM sodium hypoxanthine and 0·016 mM thymidine) at 5% haematocrit for 7 days.^25^ The albumin concentration was then reduced to 0·25%, and after a further 7 days, the media was supplemented with 10% HI human serum. On day 60, albumin was removed, and the culture transitioned to RPMI supplemented with 10% HI human serum for a further 28 days. Once the culture displayed a stable ~4-fold multiplication rate over the 32 hr life cycle (**Supplementary Figure 1**) with >60% ring-stage parasites and a parasitemia ≥3%, the adapted culture was harvested to produce the YH1-HS pre-seed bank. This pre-seed bank was cryopreserved in Glycerolyte 57 at a 1:2·2 ratio and aliquoted into 1 mL cryovials as described for other *Plasmodium* species.^26^ Cryovials were frozen at −80°C in a Thermo Scientific™ Mr. Frosty™ Freezing Container for at least 24 hours before transfer to vapour-phase liquid nitrogen (−140°C to −196°C) for storage.

**Figure 1:**
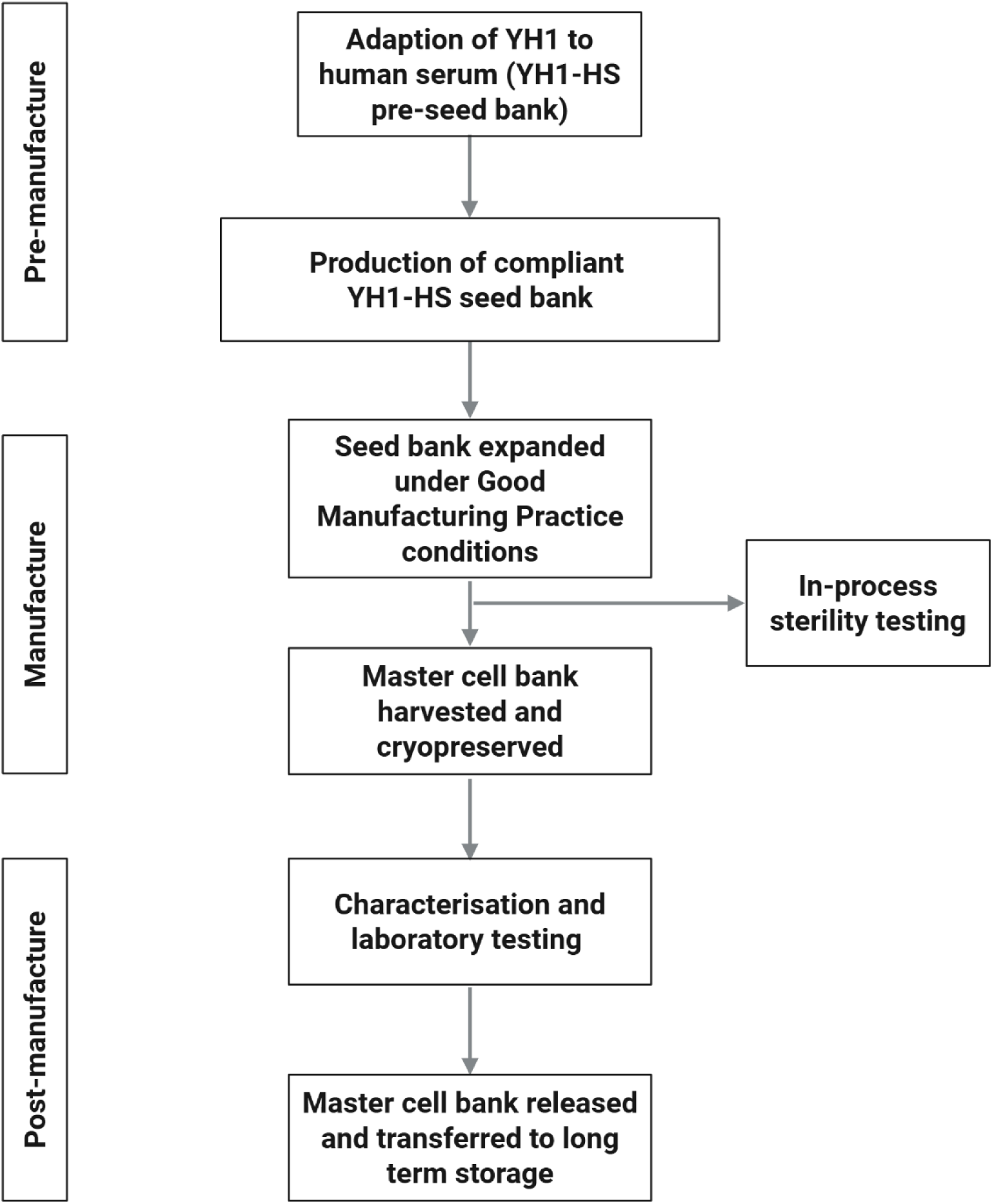
Overview of the steps involved in the production of the Good Manufacturing Practice (GMP)-compliant *Plasmodium knowlesi* YH1-HS master cell bank. Created in BioRender. Gower, J. (2025) https://BioRender.com/7k7ccot

For the production of the *P. knowlesi* YH1-HS seed bank, one vial of the pre-seed bank was thawed and the parasites cultured in RPMI 1640 supplemented with 10% HI human serum and HT supplement, at 5% haematocrit for 7 days.^25^ Once the culture contained >60% ring-stage parasites, and a parasitemia of >3%, it was cryopreserved and aliquoted into 1 mL cryovials, to create the YH1-HS seed bank. The seed bank was then stored in vapour phase liquid nitrogen until required.

### Manufacture of the *P. knowlesi* YH1-HS master cell bank using culture flasks

A single vial of the YH1-HS seed bank was thawed and culture maintained in flasks under GMP conditions^24^ at Q-Gen Cell Therapeutics (QIMR Berghofer, Brisbane), using methods previously described^25^ and materials listed in supplementary data (**Supplementary Text S1**). Cultures were sampled daily by combining all flasks, mixing, and redistributing among the flasks before gassing and returning to the incubator. Parasitemia and parasite life stages were determined from Giemsa-stained thin smears. Cultures were expanded with fresh RBCs, maintaining parasitemia at 1 – 4% until reaching ≥500 mL at ≥4% parasitemia with ≥60% ring-stage parasites. Parasitemia was then diluted to ~1% with fresh RBCs, aliquoted into 1 mL cryovials and cryopreserved as described above. The *P. knowlesi* YH1-HS parasite bank (Lot No. MBE-023) was frozen using a controlled rate freezer and stored at Q-Gen Cell Therapeutics.

### Characterisation and laboratory testing of the *P. knowlesi* YH1-HS master cell bank

To determine the *in vitro* parasite multiplication rate (PMR) of the YH1-HS parasites, one vial of parasites was thawed and cultured in standard conditions with samples taken daily for digital PCR using published primers^27,28^ (**Supplementary Table 1**). The PMR over a life cycle of 32 hours (PMR_32_) was calculated using the formula: *PMR*_32_ = 10^(1.33*m*)^, where *m* is the parasite growth rate per day estimated by the mixed-effect log-linear regression growth model (with a random intercept for culture), and 1·33 is the life-cycle of the YH1-HS strain in days. The PMR_32_ was then presented as an estimate with a 95% confidence interval (CI).

Testing of the *P. knowlesi* YH1-HS parasite bank for bloodborne infections was conducted by Pathology Queensland (Herston, Brisbane). BioReliance Ltd (Todd Campus, West of Scotland Science Park, Glasgow, G20 0XA) performed the adventitious agent screening. Sterility, endotoxin and mycoplasma testing was performed by Q-Gen Cell Therapeutics. Post-freeze parasite viability was tested using two-colour flow cytometry,^29^ while *in vitro* drug susceptibility testing against eight antimalarials was performed at Australian Defence Force Malaria and Infectious Diseases (ADFMIDI) as described previously.^30^ The results were compared to published threshold values for drug resistance.^31^

For confirmation of parasite identity, a real-time PCR method^12^ was modified to detect parasites from 10 µL of packed RBCs from the *in vitro* culture using primers and probes as above^27,28^ (**Supplementary Table 1**). DNA was extracted from parasites using the QIAamp DNA Blood kit (Qiagen, Australia).^32^ The number of *P. knowlesi* parasites in the parasite bank was quantified using a TaqMan qPCR assay targeting a 168 base pair region of the 18S rRNA gene of *P. knowlesi* **(Supplementary Text S2)**.^27,28^

For whole genome sequencing, genomic DNA was extracted from the YH1-HS seed bank and parasite bank using the NucleoSpin Blood Kit (Macherey-Nagel, Germany) with minor modifications.^33^ Sequencing was performed at the Australian Genome Research Facility (AGRF) using the NovaSeq platform to generate 150bp paired-end reads, achieving ~100 x coverage with 5 Gbp data output per sample. Whole genome sequencing and variant analysis workflows were adapted from Westaway et al^33^, while read mapping, variant discovery and genotyping were performed according to a modified version of a previously validated workflow.^34^ In brief, raw reads were quality-checked and trimmed using FastQC and Cutadapt^35^, then aligned to the PKA1-H.1 reference genome with Burrows-Wheeler Aligner (BWA).^36^ BAM files were processed using Picard (v2.26.1) and (Genome Analysis Toolkit) GATK (v3.8-1-0)^37^ for duplicate marking, base quality recalibration, and indel realignment, following Broad Institute guidelines for non-model organisms.^38^ Variants were called using GATK HaplotypeCaller and bcftools^39^, and filtered using standard quality metrics (FS≤2, MQ≥59, QD≥20). Final single nucleotide polymorphisms (SNP) and samples with >25% genotypic missingness, SNPs with minor allele frequency (MAF) < 5%, and those in hypervariable regions were also filtered using PLINK2.^40,41^

To assess population structure and genetic relatedness, we used identity-by-state (IBS) analysis and constructed neighbour-joining trees (NJT) visualised in R (version 4·3·2) using ggplot2 and ggtree.^42,43^ Principal component analysis (PCA) was performed on filtered genome-wide SNP data to investigate population clustering. Whole-genome alignments were performed using MUMmer4^44^ to compare samples with reference genomes (H-strain and YH1), and SNP concordance analysis was conducted using PLINK2 to assess pairwise similarity.

### Clinical study design and participants

The IBSM study involved four malaria-naïve healthy adults in four consecutive cohorts of one participant each. The study was conducted at the University of Sunshine Coast Clinical Trial Unit, Brisbane, Australia. Participants were eligible if they were aged 18–55 years, and met all the inclusion criteria (including a Duffy positive blood group) and none of the exclusion criteria (**Supplementary Text S3**). The primary objective was to determine the safety of the *P. knowlesi* YH1-HS malaria challenge agent. Secondary objectives included determining the *P. knowlesi* YH1-HS *in vivo* infectivity and parasite growth, and clearance profile after administration of the antimalarial drug artemether-lumefantrine.

The study utilised a dose escalation strategy. The first participant was to receive ~2,800 viable parasites, with participants in subsequent cohorts receiving a 10 or 100-fold higher dose of parasites depending on results from preceding cohorts. To prepare each dose, a vial of the *P. knowlesi* YH1-HS parasite bank was thawed, and the red cell pellet resuspended and diluted in 0·9% sodium chloride to obtain a 2 mL volume. The dilution required was calculated based on the number of RBCs and the percentage of viable parasites as determined by flow cytometry.^7^

### Study procedures

All participants were intravenously inoculated with parasite-infected RBCs on day 0. For cohorts one and two, participants visited the study site daily for the first six days for monitoring of adverse events, and blood sampling for measurement of parasitemia by qPCR targeting the gene encoding *P. knowlesi* 18S rRNA **(Supplementary Text S2)**.^27,28^ If the qPCR remained negative by day six, visits were to be reduced to three times per week; if parasites were detected, visits were to be increased to twice daily until artemether-lumefantrine (20 mg artemether / 120 mg lumefantrine; Novartis Pharmaceuticals Pty Ltd) was given, and then at specified timepoints post treatment until qPCR was negative. In cohorts three and four, the visit schedule was amended due to delayed time to parasite detection in the first two cohorts. Compulsory definitive antimalarial treatment for all participants was to be initiated when parasitaemia was ≥10,000 parasites/mL, or on day 21 (whichever occurred first); however, for cohorts three and four the protocol was amended to allow later antimalarial treatment. Artemether-lumefantrine was administered as six doses of four tablets over 60 hrs. Participant safety was evaluated by recording incidence, severity and relationship to the *P. knowlesi* YH1-HS malaria challenge agent of all adverse events as determined by self-reported symptoms, clinical examination and measurement of laboratory parameters.

### Parasite growth

The parasite growth rate following inoculation and prior to treatment with artemether-lumefantrine was estimated for participant three (the only participant with >1 positive qPCR value) using a log-linear regression model fit to the participant’s data, as implemented in R statistical package 4.3.2.^32,45^ The geometric mean of the three replicates on the log_10_ scale was used for modelling. Any data points where all replicates were negative before the first sample with detected parasites were excluded from the modelling. The PMR_32_ of the YH1-HS *in vivo* was calculated as described above for the *in vitro* PMR_32_ (only fixed effects).

### Evaluation of immune response

To evaluate the participants’ immune response to the *P. knowlesi* YH1-HS malaria challenge agent, IgG and IgM antibodies to whole parasite lysate were measured by ELISA on diluted EDTA plasma collected pre-inoculation and five additional timepoints between days eight and 34 post-inoculation (**Supplementary Text S4**).^46,47^ Optical density (OD) at 450 nm was measured using a TECAN Sunrise™ microplate reader, corrected against uninfected RBCs baseline, with two technical replicates per sample. Data were plotted using GraphPad Prism 10.1.2 (GraphPad Software, CA).

## Results

### Manufacture of the *P. knowlesi* YH1-HS master cell bank

The parasitemia of the culture as measured by microscopy throughout the adaptation period is shown in **Figure 2A**. The intraerythrocytic cycle of the YH1-HS line was 32 hours, with a stable multiplication rate of ~4-fold per cycle (**Supplementary Figure 1**). The YH1-HS seed bank was cryopreserved at 6% parasitemia, consisting of 60% rings, 30% trophozoites and 10% schizonts. 20 x 1 mL vials were aliquoted and cryopreserved. Following thawing and expansion of the seed bank over ten days of culture (**Figure 2B**), a master cell bank consisting of 182 vials of *P. knowlesi* YH1-HS-infected RBCs was successfully manufactured. At the time of harvest, the culture had a parasitemia of 4% with 78% ring-stage parasites. The harvested cells were then diluted with fresh RBCs to a parasitemia of 1·2% prior to cryopreservation. Release testing confirmed the master cell bank was suitable for use (**Tables 1 and 2**).

**Figure 2:**
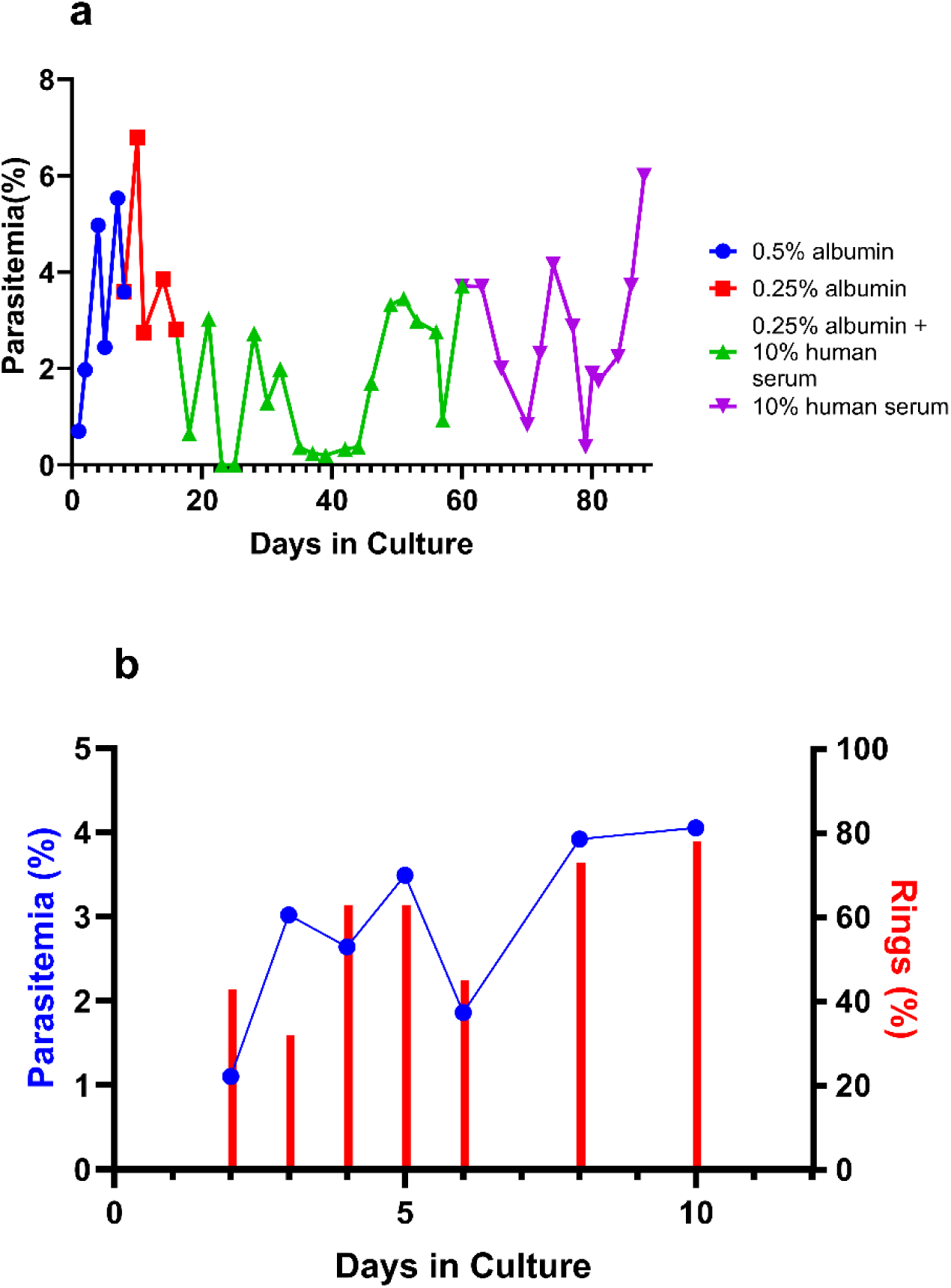
Adaption of the *P. knowlesi* YHI to growth in human serum, and growth dynamics of the YH1-HS master cell bank. **A)** Parasitemia of YH1 during the *in vitro* adaptation process to human serum, measured by microscopy over 88 days. **B)** Parasitemia and proportion of ring-stage parasites during bank manufacture of the master cell bank in flasks as determined by microscopy. Parasites showed a stable replication rate during the *in vitro* culture period.

**Table 1:** Specifications of the *P. knowlesi* YH1-HS master cell bank.

| Assay | Testing laboratory | Specifications<br>(acceptance criteria) | Results |
| --- | --- | --- | --- |
| Volume of RBC<br>pellet after thawing | QIMR Berghofer | >50 μL | Pass |
| Identification | QIMR Berghofer | Presence of *P. knowlesi* | Pass |
| Percent parasitemia | QIMR Berghofer | $\geq 0.5\%$ | 1.2% |
| % ring stage<br>parasites | QIMR Berghofer | $\geq 60\%$ | 78% |
| Viability | QIMR Berghofer | >25% | 91% |

RBC = red blood cell

**Table 2:** Laboratory testing of *P. knowlesi* YH1-HS master cell bank.

| Assay | Testing laboratory | Specifications<br>(acceptance criteria) | Results |
| --- | --- | --- | --- |
| Presence of Bovine Serum Albumin | QIMR Berghofer | <1 p.p.m. | Pass |
| Endotoxin testing | QGen | <5 EU/mL | <0.03 EU/mL |
| Sterility | QGen | No growth | Pass |
| Mycoplasma | QGen | Not Detected | Pass |
| Mycobacterium | BioReliance | Not Detected | Not Detected |
| Human immunodeficiency virus | BioReliance | Not detected | Not Detected |
| Human T-cell lymphotropic virus | BioReliance | Not detected | Not Detected |
| Epstein-Barr virus | BioReliance | Not detected | Not Detected |
| Cytomegalovirus | BioReliance | Not detected | Not Detected |
| Hepatitis C virus | BioReliance | Not detected | Not Detected |
| Hepatitis B virus | BioReliance | Not detected | Not Detected |
| Human Parvovirus(B19) | BioReliance | Not detected | Not Detected |
| Adeno-Associated virus | BioReliance | Not detected | Not Detected |
| Human herpes virus type 1, 2, 6, 7 and 8 | BioReliance | Not detected | Not Detected |
| Hepatitis A virus | BioReliance | Not detected | Not Detected |
| Simian Virus 40 | BioReliance | Not Detected | Not Detected |
| Retroviruses | BioReliance | Not Detected | Not Detected |
| Quantitative transmission electron microscopy | BioReliance | No extraneous agents observed in 200 cell profiles | No extraneous agents observed |
| Presence of viral contaminants | BioReliance | Not detected | Not Detected |
| Presence of inapparent viruses | BioReliance | Not detected | Not Detected |

### Characterisation of the *P. knowlesi* YH1-HS master cell bank

Viability of the *P. knowlesi* YH1-HS master cell bank was determined by flow cytometry from one thawed vial approximately one month after cryopreservation. The YH1-HS master cell bank contained a mean of 0·83% infected RBCs (measured in triplicate) with a mean viability of 91·24% (**Supplementary Figure 2**), exceeding the pre-defined acceptable criteria of >25% viable parasites. The concentration of *P. knowlesi* parasites in a vial of the YH1-HS parasite bank by real-time qPCR was 3·15 x 10^7^ parasites/mL (**Supplementary Table 2**). PCR assays confirmed the identity of the parasites in samples collected from day six of manufacture and day of harvest as *P. knowlesi* (**Supplementary Figure 3**). The YH1-HS parasites demonstrated susceptibility to six of the eight drugs tested (amodiaquine, lumefantrine, dihydroartemisinin, quinine, atovaquone, pyronaridine) and borderline resistance to chloroquine and mefloquine based on published values for *P. falciparum*^31^ (**Supplementary Table 3**). The *in vitro* PMR_32_ of the YH1-HS parasites was 2·47 (95% CI: 2·18-2·81).

Whole genome sequencing (WGS) was conducted on both the seed bank and master cell bank, and data compared to 152 publicly available *P. knowlesi* genomes, the majority from contemporary clinical samples.^33^ Neighbour-joining analysis based on identity by state (IBS) identified the three known genetic clusters of *P. knowlesi*, including the Peninsular-Malaysia sub-population (Peninsular) and the Malaysian-Borneo macaque associated subpopulations, *M. fascicularis* (Mf) and *M. nemestrina* (Mn).^48–50^ The YH1-HS seed bank and parasite bank samples (the “bank samples”) clustered within the Peninsular *P. knowlesi* clade (**Figure 3A**). Principal component analysis of the single nucleotide polymorphisms (SNPs) from all sequences demonstrated that the YH1-HS bank samples formed a fourth cluster separate to the Peninsula and Malaysian-Borneo associated clusters, although with three Peninsular samples (identified from the neighbour joining analysis) located within the YH1-HS cluster (**Figure 3B**). Further investigations revealed that these three Peninsular samples were laboratory-based strains, including the H strain,^51^ a Malayan strain,^52^ and the YH1 strain.^23^ The sample clustering is thus congruent with the derivation of the bank samples from both the H strain and the YH1 strain. Further analysis demonstrated >99% concordance between the bank samples and the other three strains within the same cluster in the NJT and PCA (**Supplementary Table 4**).

**Figure 3:**
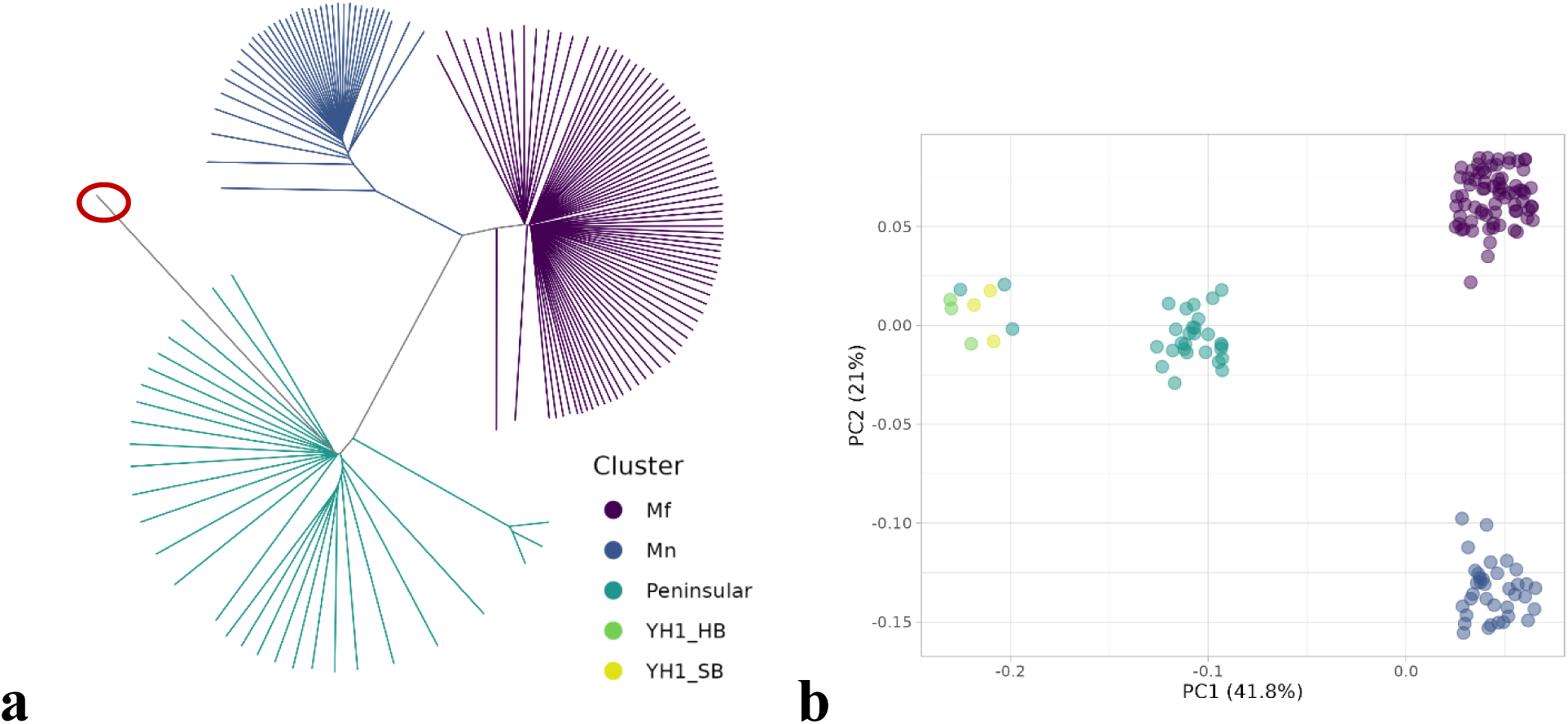
Whole Genome Sequencing of the *Plasmodium knowlesi* seed bank and master cell bank samples. **A) Unrooted neighbour-joining tree (NJT).** NJT based on identity by state (IBS) depicting three genomic clusters of *P. knowlesi* across Malaysia. Specifically, the Peninsular Malaysia sub-population (Peninsular), and Malaysian-Borneo macaque-associated subpopulations of *Macaca fascicularis* (Mf) and *M. nemestrina* (Mn). The red circle indicates the branch where the YH1-HS seed bank and master cell bank samples are located. **B) A Jittered PCA of SNP’s**. All 152 sequenced *P. knowlesi* samples and the bank samples. Four main clusters are apparent, with one cluster representing Peninsular Malaysia samples, two clusters representing the Malaysian-Borneo macaque-associated subpopulations (Mf and Mn) and a fourth cluster representing the bank samples (YH1_HB, YH1_SB) and three Peninsular Malaysia samples which are in fact laboratory-based strains from the 1960s. The three main clusters correspond exactly to the three clusters in the NJT, while the fourth cluster corresponds to the extended branch seen in the Peninsular cluster on the NJT. The first two principal components shown here account for a large portion (62·8%) of the total variation in the data. Jitter has been applied to the plot to see all samples without similar samples overlapping each other.

The mean SNP concordance between bank samples and the closest main cluster (the Peninsular cluster) was 71%, compared with a concordance of 77% within the Peninsular cluster (Supplementary Table 5). Thus, the bank samples are similar to contemporary *P. knowlesi* strains from Peninsular Malaysia (>70% concordance), but show ~6% greater genetic discordance than strains within the cluster.

### *P. knowlesi* malaria volunteer infection study

The study was conducted between January and December 2024. A total of 13 volunteers were screened for cohorts 1 – 4. Four were excluded due to not meeting the eligibility criteria, and five withdrew prior to inoculation. Four participants were enrolled (one in each cohort); all participants were inoculated, completed the study and were included in the safety analysis. Participants were aged between 20 and 51 years old, and two were female (**Table 3**). Three self-selected their ethnicity as Caucasian, and one as Hispanic/Latino.

**Table 3:** Demographics of each participant, and parasite dose administered.

| Participant | Age range,<br>years | Sex* | Race/ethnicity* | Approximate<br>number of<br>parasites<br>administered |
| --- | --- | --- | --- | --- |
| 1 | 30 - 39 | Female | Hispanic/Latino | 2,800 |
| 2 | 30 - 39 | Male | White | 28,000 |
| 3 | 20 - 29 | Female | White | 280,000 |
| 4 | 20 - 29 | Male | White | 280,000 |

\* Self-reported.

### Inoculation, parasite growth and clearance

Participant one received ~2,800 viable parasites, participant two received ~28,000 viable parasites and participants three and four both received ~280,000 viable parasites. The first two participants remained aparasitemic for 17 days post inoculation and were treated with artemether-lumefantrine on day 21 as per the study protocol. However, the second participant did have low parasitemia detected by qPCR on day 21 (~6 parasites/mL), in the sample taken immediately prior to antimalarial treatment and the results of which were not known until after treatment (**Figure 4**). For the third participant, parasitemia was first detected 17 days post-inoculation (parasite count ~3/mL), with low level parasitemia also detected on days 18 and 20 (**Figure 4**). With the participant’s consent, an urgent protocol amendment was made to allow antimalarial treatment to be administered on day 24 (the latest timepoint considered reasonable as an urgent protocol amendment) instead of day 21. Artemether-lumefantrine was therefore administered on day 24, and the participant recorded a parasitemia of 174 parasites/mL and 202 parasites/mL immediately prior to and 6 h post treatment, respectively (**Figure 4**). For participant 4, a further protocol amendment allowed for treatment to be given at day 28 if the PCR remained negative, or, if the PCR became positive, when parasitemia was >10,000 parasites/mL or at day 42 (whichever came first); however, participant four did not become parasitemic.

**Figure 4:**
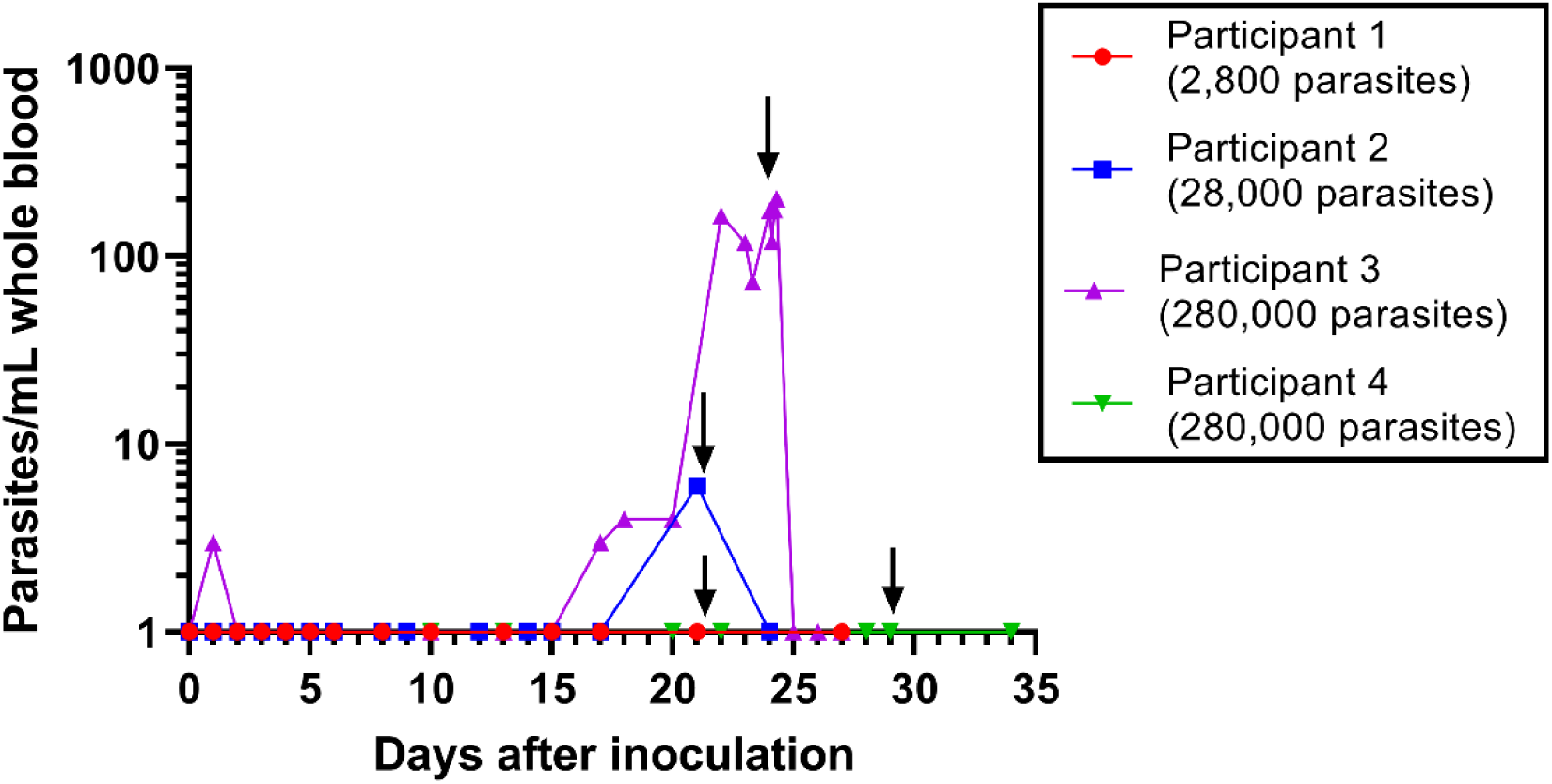
Individual participant parasitaemia-time profiles. Parasitaemia was measured using qPCR targeting the gene encoding *P. knowlesi* 18S rRNA. Vertical arrows represent the timing of antimalarial treatment initiation with artemether-lumefantrine.

For participant three, a log-linear regression model determined the growth rate estimate standard error of *P. knowlesi* YH1-HS to be 0·275 (0·056). This corresponded to a PMR_32_ of 2·33 (95% CI 1·66 – 3·25).

### Adverse events

There was a total of 22 adverse events (AEs) recorded across all four cohorts, with each participant experiencing at least one AE (**Supplementary Table 6**). As only one participant developed persistent parasitemia during the study period, the majority of AEs were not attributed to *P. knowlesi* infection (18 of 22 [82%]). Most AEs were mild (21 of 22 [95%]) with the remaining one AE of moderate severity. There were no serious adverse events (SAEs). The four AEs in Cohort 3 that were attributed to *P. knowlesi* infection were fatigue, exertional dyspnoea, hot flushes and shallow breathing. These AEs occurred between days 19 - 22, when the parasite count was increasing from ~4 parasites/mL to 165 parasites/mL (and with the participant informed of these results). Although AEs attributed to a malaria challenge agent would not normally be expected at this low level of parasitemia, the AEs were deemed possibly related to *P. knowlesi* due to the timing of the AEs and the absence of any other apparent cause. There were no clinically concerning findings from clinical laboratory analysis, ECGs, vital signs monitoring, or physical examinations.

### Immune response

Participant three generated a robust IgG response to whole *P. knowlesi* lysate on day 21 (optical density 1·0, compared to <0·5 for each of the four prior time-points) (**Figure 5**). No increase in IgM was observed for this participant at any time-point, and no IgG or IgM response was observed for the other three participants at any time-points.

**Figure 5:**
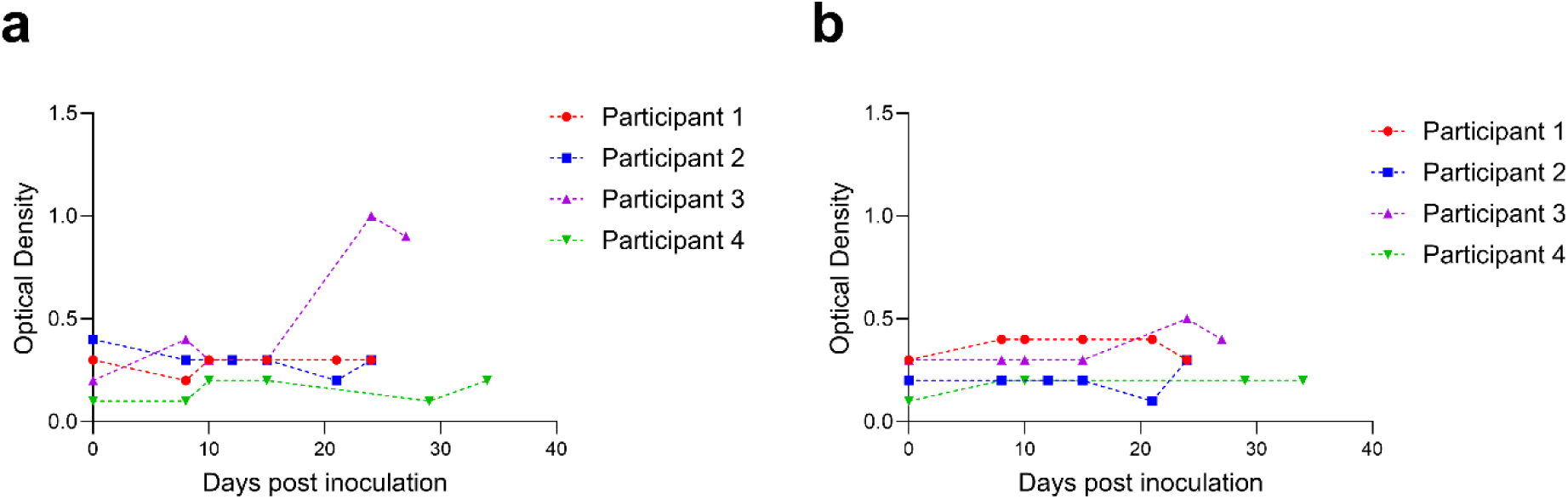
Immune response in participants following inoculation with *P. knowlesi* YH1-HS. IgG **(A)** and IgM **(B)** antibody responses to whole parasite lysate were measured longitudinally in individual participants post-inoculation. Human plasma samples were diluted 1:100 in 0·1% casein in PBS. Participant three exhibited a marked IgG response coinciding with the onset of parasitemia on Day 21.

## Discussion

The development of a reliable and reproducible *P. knowlesi* IBSM model would represent an important advance for studying this emerging zoonotic parasite. Here we report the development and characterisation of a GMP-compliant *P. knowlesi* parasite bank, and the evaluation of this bank in an IBSM study. However, while the development of a *P. knowlesi* bank suitable for use in human volunteers was successful, infectivity was suboptimal, with parasitemia detected in only one of two participants inoculated with the highest dose.

There are a number of possible explanations for the suboptimal infectivity of this *P. knowlesi* bank. First, the doses of parasites inoculated may have been insufficient to induce a reliable infection. In this study, the first participant received ~2,800 viable parasites, the second ~28,000, and the third and fourth ~280,000. The inoculum doses were selected based on experience with *P. falciparum* malaria IBSM studies,^14,22^ and with consideration of participant safety. However, in early *P. knowlesi* human infection studies conducted during the 1930s, the doses administered were often in the tens of millions of parasites (albeit not always intravenously),^18,53,54^ and incubation periods of up to 23 days were reported even despite these high doses.^18^ This suggests that the prolonged incubation period in our study (17 and 21 days in two participants) is not inconsistent with the historical data.

In addition to insufficient inoculum dose, it is possible that host factors may have contributed to reduced susceptibility to infection, particularly in the fourth participant who did not become infected despite being inoculated with the highest parasite dose. In the early *P. knowlesi* human infection studies, failure to induce infection was common, even in malaria-naive participants inoculated with highly parasitised blood.^15,53–55^ In these early studies, it is possible that Duffy-negativity may have contributed to resistance to infection in some individuals. However, it is likely that other host factors also contribute to susceptibility to *P. knowlesi* infection, and may have accounted for the variability in infection in the current study. In recent clinical studies of naturally acquired *P. knowlesi*, a wide range of parasite densities are reported, with the majority of patients having low parasitemias,^56^ and yet with some patients developing very high parasite densities with severe disease,^57^ suggesting variability in host susceptibility to parasite growth. To date, increasing age and forest-related exposure have been identified as independent risk factors for high parasitemias,^56,58,59^ although mechanisms remain uncertain. Further studies are required to investigate other host factors that may impact susceptibility to infection, such as the presence of inhibitory antibodies (including antibodies against the *P. knowlesi* Duffy binding protein alpha or *P. knowlesi* apical membrane antigen 1 required for RBC invasion),^60^ or red cell enzymopathies such as G6PD deficiency.^61^

In the current study, it is also possible that the immune evasion capabilities of the *P. knowlesi* parasites may have been impacted by long-term *in vitro* cultivation.^62^ In studies of *P. falciparum*, decreases in the transcription of erythrocyte membrane protein 1(PfEMP1)-encoding *var* genes have been demonstrated within the first 7 to10 days of culture adaptation.^63,64^ In *P. knowlesi,* the schizont-infected cell agglutination (SICA) proteins are related to and share features with PfEMP1 and the genes that encode them,^65^ and have been shown to play a role in cytoadherence of subpopulations of *P. knowlesi* to various cell types.^66,67^ It is possible that transcription of SICA proteins in the YH1-HS parasite bank may have been downregulated due to *in vitro* cultivation, possibly reducing the immune evasion capabilities and enhancing splenic clearance of the parasites.

The age of the parasites may also have contributed to the slow growth rate. The YH1-HS challenge agent was derived from the YH1 strain, which was derived from the 1965 H strain,^23^ and loss of functions may have occurred after decades of *in vitro* culture.^68^ Notably, the PMR_32_ of the *P. knowlesi* YH1-HS in this study (2·33 [95% CI 1·66 – 3.25]) was lower than that reported for *P. falciparum* (PMR_48_ 31·9 [95% CI 28·7 – 35·4])^69^ and *P. vivax* (PMR_48_ 9·9 [95% CI: 7·7–12·4]),^70^ despite the faster intraerythrocytic life cycle (32 h) of *P. knowlesi*. Although *P. knowlesi* produces fewer merozoites than *P. falciparum* and *P. vivax,*^71,72^ this is unlikely to fully account for the slow growth rate observed in the current study.

Our study had several limitations. First, the study protocol did not allow sufficient time for the third participant to develop a sustained infection. Even with a protocol amendment, we were only able to assess parasite growth over a 5-day period prior to treatment. Second, our study was limited to 4 participants, and we were therefore unable to determine whether infection would have been successful in additional participants.

In conclusion, our study demonstrated the successful development and initial evaluation of a GMP-compliant *P. knowlesi* parasite bank, suitable for use in human volunteers. However, the suboptimal infectivity and slow parasite growth highlight the need for further investigation into the adaptation, immune evasion mechanisms, and host–parasite interactions of *P. knowlesi*.

## Contributors

J.S.E.G., R.W., A.P., M.J.B., N.M.A., M.J.G., J.W. and B.E.B. contributed to the design of the study. J.S.E.G., J.M.P., D.A., H.E.J. and F.H.A. manufactured the master cell bank and prepared the challenge agent. S.M., N.S., and B.E.B. conducted the malaria volunteer infection study. N.C. and Q.C. conducted the drug sensitivity testing. D.A. and M.J.B conducted the immunological analysis. J.A.T. and C.Y.T.W. developed and conducted the qPCR assays for participant monitoring. J.A.F.W conducted the whole genome sequencing analysis. J.S.E.G, J.A.F.W., D.A., A.F.T., L.W., F.H.A., and B.E.B. analysed the data. J.S.E.G. and B.E.B. wrote the first draft of the paper. All authors approved the final draft of the manuscript. J.S.E.G. and B.E.B have accessed and verified the data.

## Data Sharing Statement

WGS data is available through NCBI: PRJNA1288158. All other data will be made available upon request through contact with the corresponding author, with an appropriate data sharing agreement in place.

## Data Availability

All data produced in the present study are available upon reasonable request to the authors.

https://www.ncbi.nlm.nih.gov/sra/PRJNA1288158

## Acknowledgements

We thank all volunteers enrolled in the malaria volunteer infection study, clinical staff at University of Sunshine Coast Clinical Trial Site, Brisbane, Australia, and Andrew Redmond for acting as a medical monitor.

The study was supported by an Australian National Health and Medical Research Council (NHMRC) Investigator Grant #2016792 to BEB and a Royal Brisbane and Women’s Hospital Foundation Grant (Perpetual Private Ltd). MJB is supported by a Snow Medical Fellowship (2022/SF167) and CSL Centenary Fellowship to MJB. MJG is supported by an NHMRC Investigator grant (#2017436). The QIMR Berghofer Clinical Malaria group receives core funding from Medicines for Malaria Venture (MMV) to support the conduct of malaria volunteer infection studies.

