## Supplementary Information for "Development of an experimental human blood-stage model for studying *Plasmodium knowlesi*"

**Supplementary Text S1: Materials used in the production of the *P. knowlesi* YH1-HS master cell bank**

RPMI-1640 media (Thermo Fisher # 22400121) was supplemented with 10% heat-inactivated (HI) pooled human serum (Australian Red Cross Lifeblood), human erythrocytes (5% haematocrit) (Australian Red Cross Lifeblood) and hypoxanthine thymidine supplement (Thermo Fisher #11067030). The erythrocytes used in the production of the *P. knowlesi* YH1-HS master cell bank were provided by the Australian Red Cross Lifeblood from a screened donor with blood group O Rh (D) Negative. The collected blood was leucodepleted at Lifeblood. The donor was screened in accordance with approved procedures and the Australian Therapeutic Goods Association regulatory requirements for donation of blood for transfusion, with further testing for additional blood-borne viruses conducted by Pathology Queensland. The pooled, HI human sera used in the production of the *P. knowlesi* YH1-HS master cell bank were collected from screened donors by Valley Biomedical (Winchester, Virginia), a USA FDA registered blood collection and processing establishment.

### Supplementary Text S2: Development of a quantitative TaqMan real-time PCR for *Plasmodium knowlesi*

To quantify the number of parasites in a vial of the master cell bank and to monitor the infection in participants, a TaqMan real-time quantitative PCR (qPCR) for *P. knowlesi* was developed. To generate a standard curve, *P. knowlesi* parasites were cultured to a high parasitemia following previously described methods.[1] Cultures were prepared for fluorescence-activated cell sorting (FACS) by staining cells at a concentration of 2 x $10^7$ red blood cells (RBCs)/mL with SYBR nucleic acid gel stain (10,000X concentrate in DMSO; Molecular Probes #S7563) diluted to 0.4 x in DPBS (Gibco #14190144). SYBR-stained cultures were sorted by FACS on two occasions using a FACS ARIA IIIu (BD Biosciences) with DIVA software Version 8 for data acquisition and sort setup. SYBR-positive cells were detected using a 488 nm laser, and logarithmic green fluorescence measured through a 530/30 band-pass filter. Uninfected RBC samples stained with SYBR were used as negative controls. A purity check was performed on sorted cells using the same settings. Sorted cells were resuspended in DPBS with parasite density assessed by counting a minimum of 2000 cells on Giemsa-stained thin films and cell concentration determined using a disposable haemocytometer. Sorted cells were stored at -80°C. DNA was extracted from sorted parasites using the QIAamp DNA Blood kit (Qiagen, Australia).[2]

Real-time qPCR was performed using a previously published TaqMan qPCR assay targeting a 168 base pair region of the 18S rRNA gene of *P. knowlesi* **(Supplementary Table 1)**.[3,4] Each qPCR reaction consisted of 12.5 µL QuantiTect Probe PCR mix (Qiagen, Australia), 0.4 µM of each primer, 0.16 µM TaqMan probe and 5 µL template DNA, in a final volume of 25 µL. Amplification and detection were performed on the Rotor-Gene Q using Rotor Gene Q Series Software, version 2.3.5 (Qiagen, Australia), in Rotor-Gene strip tubes (Qiagen, Australia). The cycling conditions were: 95 °C for 15 min for 1 cycle, followed by 45 cycles of 95 °C for 15 s and 60 °C for 1 min, with fluorescence acquisition at the 60 °C step. Oligonucleotide primers and hydrolysis probes were manufactured by Integrated DNA Technologies (Coralville, Iowa, United States). Results reported were the geometric mean of 3 replicates, with replicates with no parasites detected assigned a value of 1. This assay was accredited under ISO/IEC 17025.

**Supplementary Text S3: Clinical Study Inclusion and Exclusion Criteria**

Inclusion Criteria

Volunteers must fulfil all of the following criteria to be eligible for inclusion in this trial:

**Demographics and general considerations**

1. Healthy adults aged 18 to 55 years inclusive who will be contactable and available for the duration of the trial and up to two weeks following the EOS visit.
2. Total body weight greater than or equal to 50 kg, and a body mass index (BMI) within the range of 18 to 32 $kg/m^2$ (inclusive). BMI is an estimate of body weight adjusted for height. It is calculated by dividing the weight in kilograms by the square of the height in metres.
3. Completion of the written informed consent process prior to undertaking any trial-related procedure.
4. Must be willing and able to communicate and participate in the whole trial.
5. Agreement to adhere to Lifestyle Considerations throughout the trial duration.
6. Must be able to provide contact details of a support person (responsible adult) who is aware of the participant's participation in the study and is available to provide assistance if required (for example with contacting the participant in the event that study staff are unable to, or with transporting the participant to and from the study site if required).

**Laboratory, vital signs and ECG parameters**

7. Duffy blood group positive.
8. Vital signs at screening (measured after 5 min in the supine position):
   - Systolic blood pressure (SBP): 90–150 mmHg,
   - Diastolic blood pressure (DBP): 40–90 mmHg,
   - Heart rate (HR): 40–100 bpm.
9. At Screening and pre-inoculation with the malaria challenge agent (Day 0), 12-lead electrocardiogram (ECG) parameters after 5 minutes resting in supine position:
   - QTcF: ≤450 msec (males) or ≤470 msec (females),
   - QRS: 50–120 msec,
   - PR interval: ≤ 210 msec,
   - Normal ECG tracing unless the Principal Investigator or delegate considers an ECG tracing abnormality to be not clinically relevant.

**Contraception**

10. Women of childbearing potential (WOCBP) who anticipate being sexually active with a male during the trial must agree to use a highly effective method of birth control (see below) combined with a barrier contraceptive from the screening visit until 30 days after the end of study (covering a full menstrual cycle) and have a negative urine pregnancy test result prior to inoculation with the malaria challenge agent on Day 0.

- Highly effective birth control methods include: combined (oestrogen and progestogen containing) oral/intravaginal/transdermal/implantable hormonal contraception associated with inhibition of ovulation, progestogen-only oral/injectable/implantable hormonal contraception associated with inhibition of ovulation, intrauterine device, intrauterine hormone-releasing system, bilateral tubal occlusion, vasectomised partner, or sexual abstinence or same sex relationship.

- Female participants who are abstinent (from penile-vaginal intercourse) must agree to start a double method if they start a sexual relationship with a male during the study. Female participants must not be planning in vitro fertilisation within the required contraception period.

Women of non-childbearing potential (WONCBP) are defined as:

- Natural (spontaneous) post-menopausal defined as being amenorrhoeic for at least 12 months without an alternative medical cause with a screening follicle stimulating hormone level (FSH) >25 IU/L (or at the local laboratory levels for post-menopause).
- Premenopausal with irreversible surgical sterilization by hysterectomy and/or bilateral oophorectomy or salpingectomy at least 6 months before screening (as determined by participant medical history).

11. Males who have, or may have, female sexual partners of childbearing potential during the course of the study must agree to use a double method of contraception including condom plus diaphragm, or intrauterine device, or stable oral/transdermal/injectable/implantable hormonal contraceptive by the female partner, from the time of informed consent through to 90 days after the end of study (covering one cycle of spermatogenesis). Abstinent males must agree to start a double method if they begin a sexual relationship with a female during the study and up to 90 days after the end of study. Males that are surgically sterile, or who have undergone sterilisation and have had testing to confirm the success of the sterilisation, may also be included and will not be required to use above described methods of contraception.

Exclusion Criteria

Volunteers will be excluded from participating in the study if any of the following criteria apply:

**Medical history**

1. Known hypersensitivity to artesunate or other artemisinin derivatives, lumefantrine, proguanil/atovaquone, primaquine, or 4-aminoquinolines.
2. Any history of anaphylaxis or other severe allergic reactions, or other food or drug allergy that the Investigator considers may impact on participant safety.
3. History of convulsion (including drug or vaccine-induced episodes). A medical history of febrile convulsion during childhood (< 5 years) is not an exclusion criterion.

4. Presence of current or suspected uncontrolled chronic diseases that may impact participant safety or interpretation of clinical trial results, such as (but not limited to) cardiac or autoimmune disease, diabetes, progressive neurological disease, severe malnutrition, hepatic or renal disease, epilepsy, or asthma.
5. History of malignancy of any organ system (other than localised basal or squamous cell carcinoma of the skin or *in situ* cervical cancer), treated or untreated, within five years of screening, regardless of whether there is no evidence of local recurrence or metastases.
6. Individuals with history of schizophrenia, bipolar disorder psychoses, attempted or planned suicide, or any other severe (disabling) chronic psychiatric diagnosis including generalised anxiety disorder.
7. History of an episode of depression lasting more than 6 months that required pharmacological therapy and/or psychotherapy within the last 2 years.
8. A score of 20 or more on the Beck Depression Inventory-II (BDI-II) and/or a response of 1, 2 or 3 for item 9 of this inventory (related to suicidal ideation).

    - The BDI-II will be used as a validated tool for the assessment of depression at screening. Participants that meet criterion 8 will be referred to a general practitioner or medical specialist as appropriate. Participants with a BDI-II score of 17 to 19 may be enrolled at the discretion of the Investigator if they do not have a history of the psychiatric conditions mentioned in criterion 6 and their mental state is not considered to pose additional risk to the health of the participant during the trial or to the execution of the trial and interpretation of the data gathered.

9. History of splenectomy.
10. Cardiac/QT risk:
    - Family history of sudden death or of congenital prolongation of the QTc interval or known congenital prolongation of the QTc interval or any clinical condition known to prolong the QTc interval.
    - History of symptomatic cardiac arrhythmias or of clinically relevant bradycardia.
11. Evidence of increased cardiovascular disease risk (defined as >10%, 5-year risk for those greater than 35 years of age, as determined by the Australian Absolute Cardiovascular Disease Risk Calculator [http://www.cvdcheck.org.au/]). Risk factors include sex, age, systolic blood pressure (mm/Hg), smoking status, total and HDL cholesterol (mmol/L), and reported diabetes status.
12. Presence of clinically significant infectious disease or fever (e.g., sublingual temperature ≥38°C) within the five days prior to inoculation.

**Prior medications and treatments**

13. Any vaccination within 28 days of malaria challenge, and any vaccination planned during the study.

14. Use of prescription drugs (excluding contraceptives), investigational medical products, or non-prescription drugs or herbal supplements, that in the opinion of the investigator may potentially interfere with study interventions, within 14 days or five half-lives (whichever is longer) prior to malaria challenge.
15. Individual who has ever received a blood transfusion.

**Malaria exposure**

16. Any history of malaria or participation in a previous malaria challenge trial or malaria vaccine trial.
17. Must not have had malaria exposure that is considered by the Principal Investigator or their delegate to be significant. This includes but is not limited to: history of having travelled to or lived (>2 weeks) in a malaria-endemic region during the past 12 months or planned travel to a malaria-endemic region during the course of the trial; history of having lived for >1 year in a malaria-endemic region in the past 10 years; history of having ever lived in a malaria-endemic region for more than 10 years inclusive. For endemic regions see https://malariaatlas.org/explorer/#/, Bali is not considered a malaria-endemic region.

**Alcohol use and smoking**

18. History or presence of alcohol abuse (regular alcohol consumption in males >21 units per week and females >14 units per week (1 unit = ½ pint beer, or a 25 mL shot of 40% spirit, 1.5 to 2 units = 125 mL glass of wine, depending on type), or drug habituation, or any prior intravenous usage of an illicit substance.
19. Any individual who currently smokes cigarettes on a daily basis (including e-cigarettes, vaping, and other nicotine use).

**Blood donation**

20. Blood product donation to any blood bank during the 8 weeks (whole blood) or 4 weeks (plasma and platelets) prior to malaria challenge.
21. Individual unwilling to defer blood donations for at least twelve months after the EOS visit.

**Laboratory results**

22. Haematology, biochemistry or urinalysis results at screening or at the eligibility visit (Day -1 to Day -3) that are outside of the standard clinically acceptable laboratory ranges and are considered clinically significant by the Principal Investigator.
23. Positive result for: hepatitis B surface antigen (HBs Ag), anti-hepatitis C virus (anti-HCV) antibodies, anti-human immunodeficiency virus 1 and 2 antibodies (anti-HIV1 and anti-HIV2 Ab), COVID-19 by rapid test on Day 0, red blood cell alloantibodies.

24. Positive urine drug test. Any drug listed in the urine drug screen unless there is an explanation acceptable to the Investigator (e.g., the participant has stated in advance that they consumed a prescription or over-the-counter product that contained the detected drug) and the participant has a negative urine drug screen on retest by the pathology laboratory.
25. Positive alcohol breath test.
26. Positive serum pregnancy test at screening visit, positive urine pregnancy test on Day 0.

**Other**

27. Individual who, in the judgement of the Investigator, is likely to be non-compliant during the trial
28. Individual who is an Investigator, research assistant, pharmacist, trial coordinator, or other staff thereof, directly involved in conducting the trial.
29. Individual without good peripheral venous access.
30. Individual who is breastfeeding or lactating.

**Supplementary Text S4: Methodology for determining antibody mediated immune response to *P. knowlesi* YH1-HS infection**

*Plasmodium knowlesi* YH1 was cultured in O+ human red blood cells (RBCs) at 5% haematocrit in RPMI 1640 media (Thermo Fisher #31800089) supplemented with 0·2% sodium bicarbonate, 25 mM HEPES, 31.25 µg/mL Gentamicin (Thermo Fisher #15750060), 367 µM hypoxanthine (Merck #H9636) and 0·5% AlbuMAX™ II Lipid-Rich BSA (Thermo Fisher #11021045). Parasite infected RBCs (pRBCs) were cultured to 10% parasitemia at late trophozoite stage before enrichment to 95% purity by magnetic separation with CS column (Miltenyi #130-041-305). pRBCs were lysed by repeatedly freeze/thawing in dry ice/ethanol slurry and 37 °C water bath. Samples were sonicated then centrifuged at 10,000 x g. Supernatant was collected and stored at -80 °C. This process was repeated with uninfected RBCs (uRBCs) which were used as control lysate.

Parasitised RBC and uRBC lysate was thawed and coated separately at 1:3,000 in PBS onto 96-well flat bottom Nunc MaxiSorp® plates (Thermo Fisher #44-2404-21), then incubated at 4 °C overnight. Plates were blocked with 1% casein (Merck #C8654-500G) in PBS for 2 h at 37 °C, before washing 3x with PBS + 0.05% Tween-20 (PBS-T). Human plasma samples were diluted 1:100 in 0·1% casein in PBS and incubated in plate for 1 h at room temperature. Bound IgG and IgM were captured separately with mouse anti-human IgG (Thermo Fisher #M110303, 1:1,000 in 0·1% casein in PBS) and mouse anti-human IgM (Thermo Fisher #054900, 1:1,000 in 0·1% casein in PBS). Mouse antibody was detected with 1:1,000 goat anti-mouse IgG HRP conjugate (Thermo Fisher #A16066) in 0·1% casein in PBS for 1 h at room temperature. ELISA plates were washed 3x with PBS-T after each antibody incubation. TMB Chromogen Solution (Thermo Fisher #002023) was added for 10 min to visualise HRP, and reaction was stopped using 1 M hydrochloric acid. Optical density (O.D.) at 450 nm (absorbance wavelength) and 620 nm (reference wavelength) was determined by reading plates on TECAN Sunrise™ microplate reader. Reference wavelength reading was subtracted from absorbance wavelength reading for each well, before uRBC value was removed from pRBC value for each sample. Technical replicates were performed in separate plates and averaged for each sample.

**Supplementary Table 1: Primers and probes used for real-time PCR for the detection of *P. knowlesi***

| Primer/Probe | Sequence | Ref |
|---|---|---|
| Forward Primer (Plasmo1) | 5’-GTTAAGGGAGTGAAGACGATCAGA-3’ | 3,4 |
| Reverse Primer (Plasmo2) | 5’-TTATGAGAAATCAAAGTCTTTGGGTT-3’ | |
| Plasprobe_*Pk* | 5’-FAM-CTCTCCGGAGATTAGAACTCTTAGATTGCT-BHQ1-3’ | |

**Supplementary Table 2: Summary of results obtained from real-time qPCR**

| Subject | EHV CP | CP of duplicates | Calculated (p/mL) | Average (p/mL) | Adjusted (p/mL) for dilution (replicate) | Adjusted (p/mL) for dilution (sample) |
|---|---|---|---|---|---|---|
| MBE-023, Exp 1 (1:10,000) | 24·18 | 27·65 | 3,598 | 3,489 | 34,893,333 | 31,503,333 |
| | | 27·77 | 3,314 | | | |
| | | 27·67 | 3,556 | | | |
| MBE-023, Exp 2 (1:10,000) | 24·36 | 28·09 | 2,651 | 2,811 | 28,113,333 | |
| | | 27·97 | 2,879 | | | |
| | | 27·96 | 2,904 | | | |

Exp = experiment, EHV = equine herpes virus (control); p/mL = parasites per mL; CP = crossing point

**Supplementary Table 3: *In vitro* drug susceptibility profile of the YH1-HS master cell bank**

| Antimalarial Drug | Cut-off value $IC_{50}$ (nM) | $IC_{50}$ (nM) | Susceptibility profile |
|---|---|---|---|
| Amodiaquine | 80 | 9·40 | Sensitive |
| Lumefantrine* | | 18.1 | Sensitive |
| Chloroquine | 80 - 100 | 30·7 | Resistant |
| Dihydroartemisinin* | | 1·70 | Sensitive |
| Mefloquine | 20 – 30 | 18·1 | Resistant |
| Quinine | 300 - 500 | 146·9 | Sensitive |
| Atovaquone* | | 4·21 | Sensitive |
| Pyronaridine* | | 1·80 | Sensitive |

*No established threshold values

**Supplementary Table 4: Summary of SNPs concordance rates between the bank samples and clustered NJT and PCA samples.**

| Challenge Agent Sample | Clustered sample | Cluster sample alias | Average SNPs interrogated | Average SNPs difference between genomes | Average concordance (%) |
|---|---|---|---|---|---|
| YH1-HS | SRR2222335 | H strain | 513,862 | 44 | 99·99137252 |
| seed bank | SRR2225467 | Malayan strain | 513,843 | 51 | 99·99007479 |
| | SRR3135172 | YH1 strain | 513,869 | 37 | 99·99312406 |
| YH1-HS | SRR2222335 | H strain | 513,864 | 48 | 99·99065902 |
| master cell bank | SRR2225467 | Malayan strain | 513,846 | 53 | 99·98962075 |
| | SRR3135172 | YH1 strain | 513,867 | 36 | 99·99292943 |

**Supplementary Table 5: Summary of average SNPs concordance rates between the bank samples and Peninsular cluster as well as average concordance within the Peninsular cluster.**

| Pairwise | Average SNPs interrogated | Average SNPs different between genomes | Average concordance (%) |
|---|---|---|---|
| Seed bank - Peninsular | 511,407 | 147,654 | 71·12782639 |
| MCB - Peninsular | 511,408 | 147,657 | 71·12746507 |
| Peninsular - Peninsular | 510,270 | 116,222 | 77·22339435 |
| Peninsular - Mf | 509,935 | 196,272 | 61·5141894 |
| Peninsular - Mn | 509,607 | 199,078 | 60·93492631 |

MCB = master cell bank

**Supplementary Table 6: Adverse events recorded for each participant**

| Adverse Event | Participant 1 | Participant 2 | Participant 3 | Participant 4 |
|---|---|---|---|---|
| Headache | 4 | 3 | 1 | 1 |
| Lower back ache (myalgia) | 1 | 0 | 0 | 0 |
| Light headedness | 1 | 0 | 0 | 0 |
| Covid-19 infection | 1 | 0 | 0 | 0 |
| Myalgia | 1 | 0 | 0 | 0 |
| Left shoulder tendonitis | 0 | 1 | 0 | 0 |
| Upper limb fatigue | 0 | 0 | 1 | 0 |
| Upper limb myalgia | 0 | 0 | 1 | 0 |
| Fatigue | 0 | 0 | 1 | 0 |
| Exertional dyspnoea | 0 | 0 | 1 | 0 |
| Hot flush | 0 | 0 | 1 | 0 |
| Shallow breathing | 0 | 0 | 1 | 0 |
| Sinus congestion | 0 | 0 | 1 | 0 |
| Bruising left cubital fossa | 0 | 0 | 1 | 0 |
| **Total** | **8** | **4** | **9** | **1** |

**Supplementary Figure 1: A) Parasitaemia of *P. knowlesi* YH1-HS cultured *in vitro* over time, measured by microscopy to assess growth kinetics.** Both replicates show an increase in parasitemia at 32 hours (dotted line), indicating the reinvasion of a new cycle of parasites. **B) Proportion of asexual life stages (rings, trophozoites, schizonts) at indicated time points, based on morphological assessment by microscopy.**

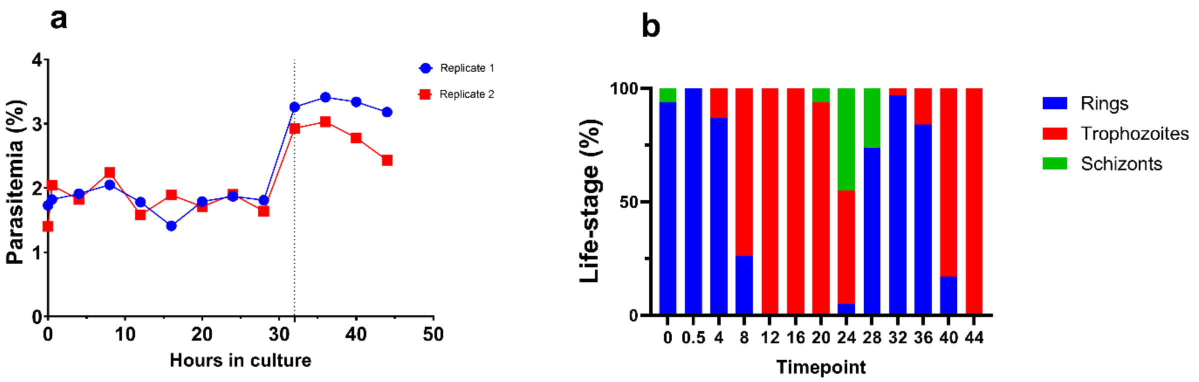

**Supplementary Figure 2:** YH1-HS master cell bank viability testing. **A)** Representative gating strategy, quadrant and region set up for *P. knowlesi* YH1-HS master cell bank MBE023 Tube A (1:41 dilution) and uninfected red blood cells single stain control samples post freeze. **B)** Flow cytometric determination of parasite viability in *P. knowlesi* YH1-HS master cell bank MBE023 Tube A (1:41 dilution).

**a**

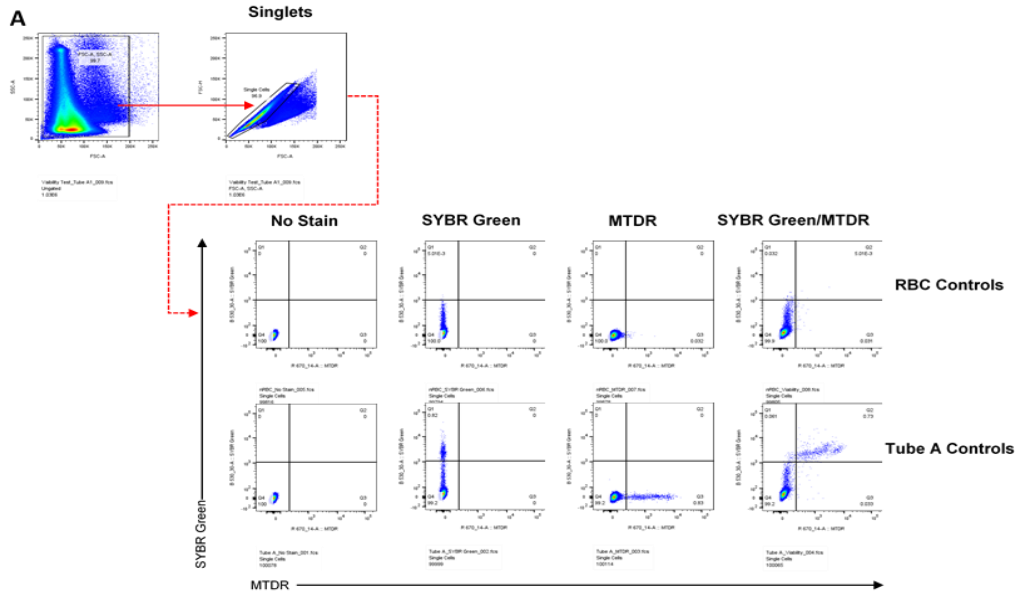


**b**

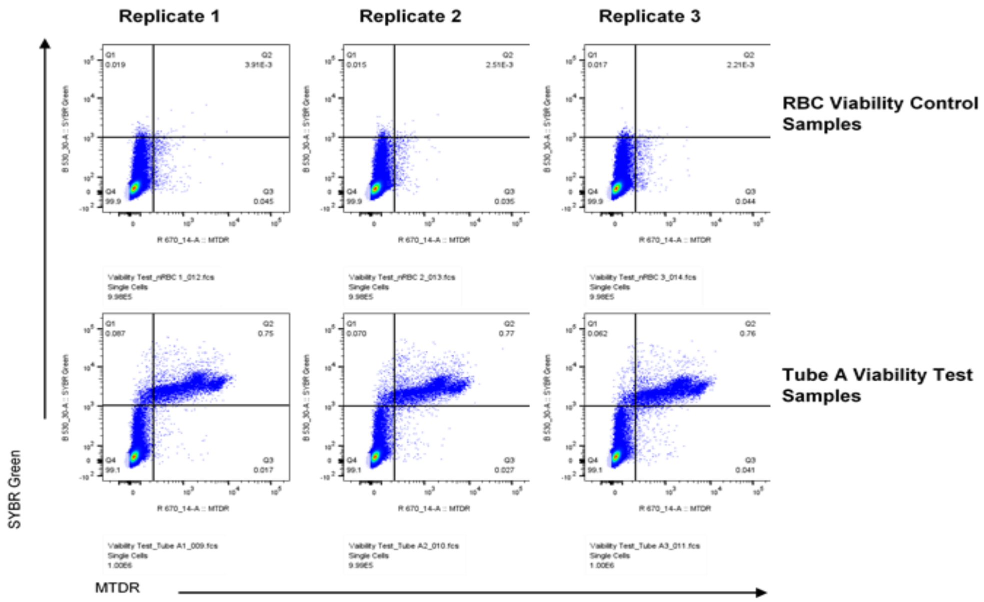

**Supplementary Figure 3: YH1-HS master cell bank identity testing**. Quantification cycle (Cq) values as determined by qPCR on duplicate samples taken during banking process (Day 6 and harvest). Samples were run in triplicate with error bars displaying geometric mean and geometric standard deviation.

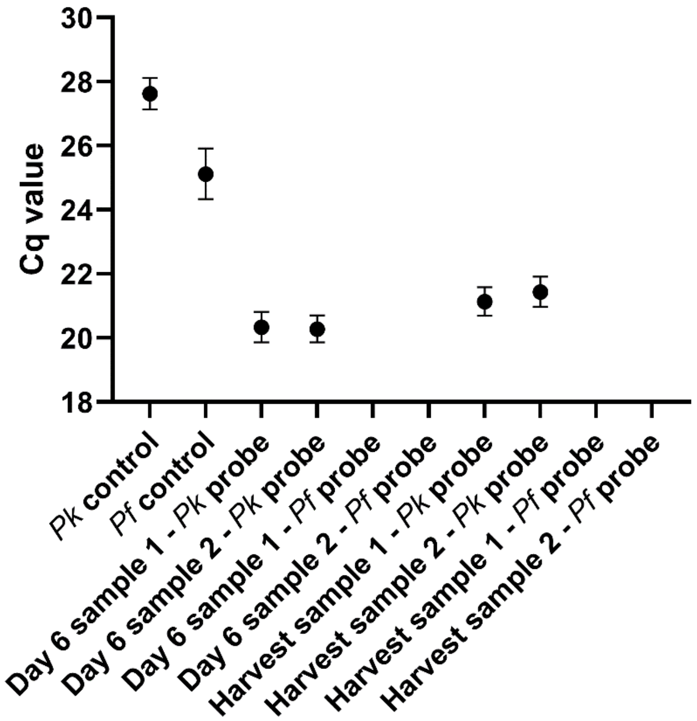
